# Economic Crisis and Mental Health during the COVID-19 Pandemic in Japan

**DOI:** 10.1101/2021.03.20.21254038

**Authors:** Tetsuya Matsubayashi, Yumi Ishikawa, Michiko Ueda

## Abstract

**Background:** The economic crisis induced by the COVID-19 pandemic can have a serious impact on population mental health. This study seeks to understand whether the economic shocks associated with the pandemic have a differential impact by sex because the current pandemic may have disproportionally affected women compared to men.

**Methods:** We analyzed data from original online monthly surveys of the general population in Japan conducted between April 2020 and February 2021 (N=9000). We investigate whether individuals who had experienced a major job-related were more likely to have experienced depressive symptoms (PHQ-9) and anxiety disorders (GAD-7) during the pandemic and also if its effect varied by sex.

**Results:** The results of logistic regression suggest that depressive and anxiety symptoms were more prevalent among those who had recently experienced drastic changes in employment and working conditions, as well as among individuals with low income and those without college education. We also found that female respondents who had experienced a major employment-related change were more likely to have experienced both depression and anxiety disorders, but its effect on male workers was limited to depressive symptoms.

**Limitations:** We do not have data on the pre-COVID mental health conditions of our respondents, and our findings are descriptive. Some segments of the population may not be represented in our sample because our surveys were conducted online.

**Conclusions:** COVID-induced economic shocks can have a differential detrimental effect on mental health depending on the sex of workers. The mental health of female workers can be particularly vulnerable.

## Introduction

The COVID-19 pandemic has triggered a major economic crisis in many parts of the world. International Monetary Fund reported that the global economic growth in 2020 fell by −3.5 percent due to the pandemic (IMF 2021). This is the worst economic downturn since the Great Depression of the 1930s, being far worse than the 2008 Global Financial Crisis.

The findings of past studies suggest that mental health conditions tend to deteriorate in times of economic downturns (e.g., Bradford and Lastrapes 2014; Barr, Kinderman, and Whitehead 2015; Johnston, Shields and Suziedelyte 2020; Wang and Fattore 2020). Thus, the economic crisis induced by the COVID-19 pandemic can have a serious impact on population mental health. Recent studies have found that individuals who have experienced COVID-induced economic shocks, such as reduced workload and income loss, were more likely to have worse mental health conditions (See a review by Xiong et al 2020). The negative effects of economic shocks on psychological well-being have been shown to be larger among economically vulnerable individuals (Cheng et al. 2021; Witteveen and Velthorst, 2020).

This study seeks to understand whether the economic shocks associated with the pandemic have a differential impact by sex. It is possible that the current pandemic has disproportionally affected women compared to men; it affected industries that were more likely to be served by women, including tourism and food services (IMF 2020). In addition, the burden of unpaid domestic care tends to be borne by women, and some women might have been forced to leave the workforce to care for their family members in response to school closure and other prevention measures. These changes can make them economically vulnerable, particularly because women are less likely to have a position that provides social protection (ILO 2020), which can negatively impact their psychological status.

To understand the effects of COVID-related economic consequences on the psychological well-being of men and women, we analyzed data from original online monthly surveys of the general population in Japan conducted between April 2020 and February 2021. In addition to testing the possibility of differential effects of economic shocks on psychological distress by sex, our study has two distinctive improvements over previous studies. First, our data include a total of 9,000 respondents who are nationally representative of the Japanese population. The sample size is much larger than that of previous studies, with the exception of Cheng et al. (2021). Second, in contrast to previous studies focusing on the early phase of the pandemic (Cheng et al. 2021; Witteveen and Velthorst 2020), the data of our study come from multiple phases of the pandemic, covering a much longer period. This feature allows us to obtain up-to-date and more generalizable evidence.

## Methods

Our analysis used data from a series of monthly online surveys that we have been conducting since April 2020. We delegated the survey to one of the major commercial survey companies in Japan. Each month, the company sent out screening questions to approximately 10,000 registered individuals and then constructed a final sample of 1,000 respondents so that they were representative of the Japanese population in terms of sex, age groups, and areas of residence. This study used the surveys taken between June 2020 and February 2021 (N=9,000), as the question on their economic conditions were asked only since June.

As measures of population mental health, we used the Patient Health Questionnaire nine-item scale (Spitzer et al. 1999) for depressive symptoms and the seven-item General Anxiety Disorder Scale (GAD-7) (Spitzer et al. 2006) for anxiety disorders. The Cronbach’s alpha value for each of the scales was 0.91 and 0.92, respectively. For our regression analysis, we generated an indicator variable that equaled one if the total score of the PHQ-9 was 10 or higher for depressive symptoms and one if the total score of the GAD-7 was 10 or higher for anxiety symptoms.

To measure the experience of job-related adverse changes during the pandemic, we asked the respondents whether they had experienced a job loss, layoff, or significant reduction in working hours in the previous three months. We categorized the respondents into three groups: “adverse change,” “no change,” and “not in the labor force” (the reference group).

To account for their socioeconomic status, we also asked about their household income in the previous year, educational attainment, and employment status/type of employment. The respondents were categorized into four income groups: “low income” (<4 million yen), “middle income” (between 4 to 8 million yen), “high income” (>8 million yen; the reference), and “income unknown.” Their educational attainment was captured by an indicator variable that equaled one if they had no college degree. Thus, those with a college degree were the reference group. We also categorized the respondents into five groups according to their employment status and type of employment: working regularly for a permanent position (the reference), working regularly for a non-permanent position, self-employed, searching for a job, and not in the labor force.

We conducted logistic regression in which the prevalence of depressive and anxiety symptoms was regressed on the indicator variables for the presence of an adverse job-related change, socioeconomic status, indicator variables for their age group (<40 years old, between 40 and 64 years of age, and aged 65 years or older), and survey months for all respondents and by sex. In addition, we controlled for local pandemic situations using the (logged) monthly number of new Covid-19 cases and deaths in the prefecture where the respondents resided at the time of the survey. Estimation results were transformed into odds ratios and presented with a 95% confidence interval using the forester library in R (Boyes 2021).

The survey was approved by the Ethics Review Committee on Human Research of Waseda University (approval #: 2020-050).

## Results

Table 1 reports the number and percentage of respondents in each category of the economic demographic variables for all respondents and by sex. About 12% of the respondents experienced an adverse change in employment status during the study period. About one-third of the respondents were categorized into low- or middle-income groups. Half of the respondents had no college degree. Finally, about 11% of the respondents reported working in a non-permanent position, while about 5% of them had been searching for a job. We anticipate that these groups were most likely to be affected by the economic crisis and thus likely to show a higher prevalence of depressive and anxiety symptoms.

**Table 1:**
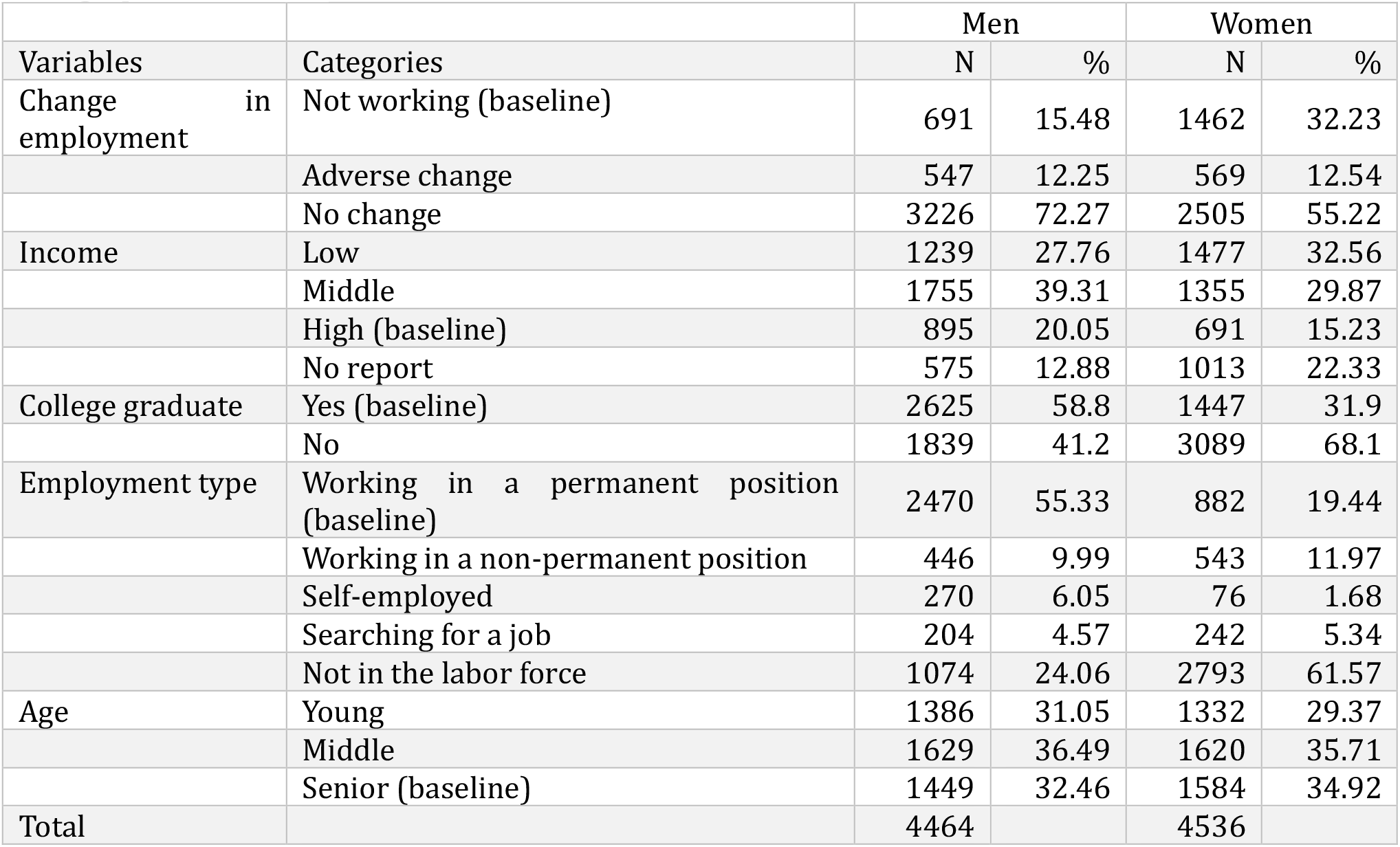
The number and percentage of respondents in each category of the economic and demographic variables

Figure 1 presents the estimation results when the PHQ-9 was used as the outcome variable (left panel) and the GAD-7 as the outcome (right panel). The indicator variables for sex (when we used all respondents), age group, survey months, and the monthly total number of new cases and deaths in each prefecture were always included in the estimation, but the results are not reported.

**Figure 1:**
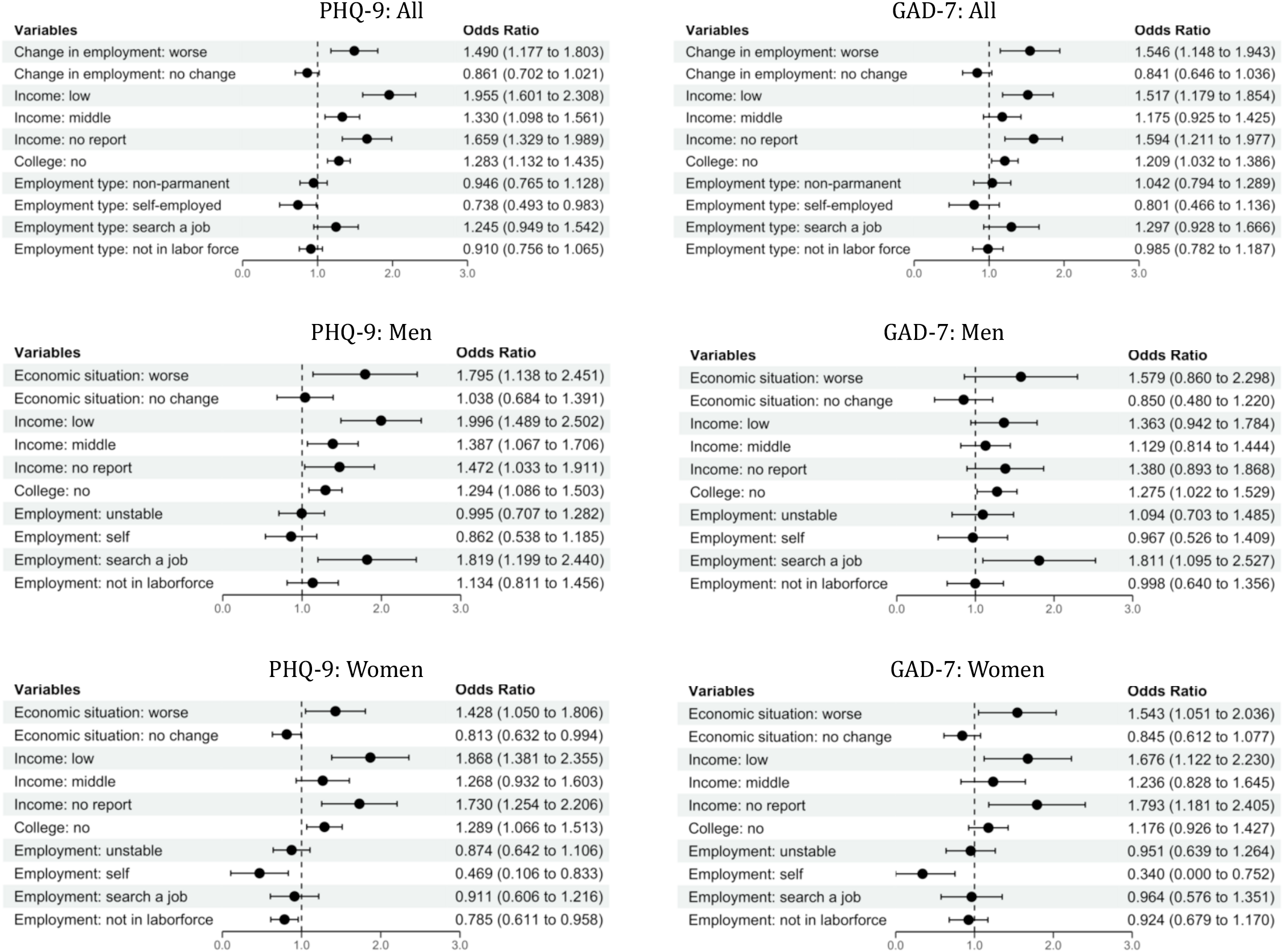
Estimated relationships between personal economic situations and the prevalence of depressive symptoms and anxiety. Note: Odds ratios ware based on logistic regression estimates. Sex, age groups, survey rounds, and the (log) number of monthly total of COVID-19 infections and deaths in the prefectures were adjusted.

The top panel contains the results when all the respondents were included in the estimation. Those who had experienced a major job-related change exhibited a higher prevalence of depressive and anxiety symptoms by about 50% in the odds (1.490, 95% CI: 1.177-1.803, 1.546, 95% CI: 1.148-1.943, respectively), compared to those who were not in the labor force. Relative to those with high income, those with low income also had higher odds of having depressive and anxiety symptoms (1.955, 95% CI: 1.601-2.308, and 1.517, 95% CI: 1.179-1.854, respectively) than those with high income. Those with middle income and those who did not report their income also exhibited a higher prevalence of depressive symptoms. Those without college education also showed a higher prevalence of depressive (1.283, 95% CI: 1.132-1.435) and anxiety symptoms (1.209, 95% CI: 1.032-1.386) than those with college education. We found no difference in prevalence across employment status and type.

According to the results stratified by sex, female respondents who had experienced a major change in their employment status or working conditions were more likely to have experienced depressive (1.428, 95% CI: 1.050-1.806) and anxiety symptoms (1.543, 95%: 1.051-2.036) compared to those who were not in the labor force. We also found that female workers who were unemployed had higher odds of developing depressive and anxiety symptoms. Similarly, women with low income or those who refused to answer their income level were more likely to suffer from depressive and anxiety symptoms compared to their high-income counterparts.

Male workers who had experienced a job or income loss were more likely to suffer from depression (1.795, 95% CI: 1.138-2.451) relative to those who were not in the labor force. In addition, compared to those with high income, those who had lower income levels or those who did not report their income level had higher odds of depressive and anxiety symptoms. Low-income males were twice as likely to have depressive symptoms compared to their high-income counterparts. In addition, men who were looking for a job had a high prevalence of depressive and anxiety symptoms.

## Discussion

This study examined whether individuals who had experienced a major job-related change as well as those with low socioeconomic status were more likely to have experienced depressive and anxiety symptoms during the COVID-19 pandemic. We found that depressive and anxiety symptoms were more prevalent among those who had recently experienced drastic changes in employment and working conditions, as well as among individuals with low income and those without college education. Thus, our overall findings suggest that economically vulnerable individuals have unfavorable mental health conditions during the pandemic.

We also found that the adverse effects of employment status change on psychological well-being differed between the sexes. We found that female respondents who had experienced a major employment-related change were more likely to have experienced both depression and anxiety disorders, but its effect on male workers was limited to depressive symptoms. Male workers who were unemployed and looking for a position were more likely to suffer from psychological distress, whereas we did not find such a pattern among female workers.

This study contributes to the literature by documenting the differential effects of COVID-induced economic shocks on the mental health of women and men. Our overall findings are consistent with recent studies that demonstrated the negative impact of economic shocks on mental health conditions during the pandemic (Dawel et al. 2020; Witteveen and Velthorst 2020), but none of these studies investigated whether the psychological impact of work-related changes differed by sex.

The observed differences between sexes may be attributable to the fact that the current pandemic puts disproportionate burdens on women in multiple domains, which can make them vulnerable both economically and psychologically. Women were more likely to experience job losses as a result of the pandemic because they were more likely to work in the affected industries, including tourism and food industry (McKinsey 2020). In addition, some women faced increased childcare responsibilities due to long-term school and daycare closures, which may have forced them to stop looking for a job. Thus, the COVID-19 pandemic caused some women multifaceted challenges, which might explain why economically vulnerable women were more likely to suffer from both depression and anxiety disorders.

This study has several limitations. We do not have data on the pre-COVID mental health conditions of our respondents, and thus our findings are descriptive, not causal. Additionally, although our surveys were designed so that our respondents were representative of the Japanese population in terms of their sex, age group, and area of residence, it is possible that some segments of the population were not represented in our sample because they were online surveys.

Despite these limitations, this study presented evidence that COVID-induced economic shocks can have a differential detrimental effect on mental health depending on the sex of workers. The economic ramifications of the COVID-19 pandemic can remain with us much longer than the disease itself. More work is necessary to identify vulnerable individuals during the pandemic so that we can monitor their mental health status.

## Data Availability

Due to the sensitive nature of the data, we cannot share the data.

## References

Barr, B., Kinderman, P., & Whitehead, M. (2015). Trends in mental health inequalities in England during a period of recession, austerity and welfare reform 2004 to 2013. Social Science & Medicine, 147, 324–331.

Bradford, W. D., & Lastrapes, W. D. (2014). A prescription for unemployment? Recessions and the demand for mental health drugs. Health economics, 23(11), 1301-1325.

Boyes, R. (2021). An R package for creating publication-ready forest plots. https://github.com/rdboyes/forester

Cheng, Z., Mendolia, S., Paloyo, A. R., Savage, D. A., & Tani, M. (2021). Working parents, financial insecurity, and childcare: mental health in the time of COVID-19 in the UK. Review of Economics of the Household, 1–22.

International Labor Organization. (2020). COVID-19 and the world of work: Impact and policy responses. https://www.ilo.org/wcmsp5/groups/public/---dgreports/---dcomm/documents/briefingnote/wcms_738753.pdf

International Monetary Fund. (2021). World Economic Outlook Update. Retrieved from https://www.imf.org/en/Publications/WEO/Issues/2021/01/26/2021-world-economic-outlook-update

Johnston, D. W., Shields, M. A., & Suziedelyte, A. (2020). Macroeconomic Shocks, Job Security, and Health: Evidence from the Mining Industry. American Journal of Health Economics, 6(3), 348-371.

Leske, S., Kõlves, K., Crompton, D., Arensman, E., & De Leo, D. (2021). Real-time suicide mortality data from police reports in Queensland, Australia, during the COVID-19 pandemic: an interrupted time-series analysis. The Lancet Psychiatry, 8(1), 58-63.

McKinsey & Company. (2020). COVID-19 and gender equality: Countering the regressive effects. https://www.mckinsey.com/featured-insights/future-of-work/covid-19-and-gender-equality-countering-the-regressive-effects

Spitzer RL, Kroenke K, Williams JB. Validation and utility of a self-report version of PRIME-MD: the PHQ primary care study. Primary Care Evaluation of Mental Disorders. Patient Health Questionnaire. JAMA. 1999 Nov 10;282(18):1737–44.

Spitzer RL, Kroenke K, Williams JBW, Löwe B. A brief measure for assessing generalized anxiety disorder: the GAD-7. Arch Intern Med. 2006 May 22;166(10):1092–7.

Xiong J, Lipsitz O, Nasri F, Lui LMW, Gill H, Phan L, Chen-Li D, Iacobucci M, Ho R, Majeed A, McIntyre RS. Impact of COVID-19 pandemic on mental health in the general population: A systematic review. J Affect Disord. 2020 Dec 1;277:55–64.

Wang, Y., & Fattore, G. (2020). The impact of the great economic crisis on mental health care in Italy. The European Journal of Health Economics, 21(8), 1259-1272.

Witteveen, D., & Velthorst, E. (2020). Economic hardship and mental health complaints during COVID-19. Proceedings of the National Academy of Sciences, 117(44), 27277–27284.

